# Chronic Disease and Risk Factor Prevalence in Multiracial Subgroups: California, 2014–2023

**DOI:** 10.1101/2025.06.19.25329941

**Authors:** Tracy Lam-Hine, Michelle C. Odden, Aliya Saperstein, Tainayah W. Thomas, David H. Rehkopf

## Abstract

**Background:** Multiracial adults represent a growing U.S. population but are often grouped together or reassigned to single-race categories in public health data. Aggregation can obscure important variation across subgroups, limiting opportunities for targeted prevention.

**Methods:** We analyzed 2014–2023 California Behavioral Risk Factor Surveillance System data (n=100,177) to estimate prevalence of 28 health indicators across racial and ethnic groups, including disaggregated Multiracial subgroups. We categorized participants based on all self-identified races and aggregated subgroups with N<50. We standardized prevalence by age and sex using 2020 California census data, calculated relative standard errors, and used survey-weighted methods to compare prevalence and subgroup differences.

**Results:** Among 100,177 participants, Multiracial subgroups had the highest prevalence for 24 of 28 outcomes. American Indian or Alaska Native–Black and Hispanic–Black–White adults had the highest prevalence of chronic conditions, poor general health, and disability. In contrast, Asian Multiracial subgroups (e.g., Asian–Black, Asian–Pacific Islander) more often had the lowest prevalence, though Asian–White adults were not consistently the healthiest subgroup. Differences across Multiracial subgroups exceeded 20 percentage points for nearly half of all outcomes.

**Discussion:** Wide health variation among Multiracial adults is masked by common aggregation practices. Subgroups with the highest burden may be overlooked if data are not routinely disaggregated. Public health surveillance systems should expand capacity to collect and report disaggregated race and ethnicity data to better inform prevention strategies.

## Introduction

One in ten U.S. residents (34 million) identified as Multiracial (two or more races) in 2020 (*1*). As a group, Multiracial adults have higher national prevalence of several chronic and mental health conditions compared to monoracial (single-race) adults (*2–4*). Like all social categories, “Multiracial” includes heterogeneous subgroups with varying health and socioeconomic profiles (*5*). Disaggregation enables subgroup prioritization for targeted prevention (*6*), but sample size limitations and lack of consensus on how to disaggregate the Multiracial category have hindered such analyses (*7*). This cross-sectional study used a population-representative dataset to estimate prevalence of 28 health indicators, disaggregating the Multiracial category alongside other racial and ethnic groups.

## Methods

The Behavioral Risk Factor Surveillance System (BRFSS) is a phone-based survey conducted by state health departments to monitor health issues among U.S. adults (18+). Data on each racial and ethnic group participants selected are unavailable in the national public use BRFSS. Instead, we requested these data directly from the California Department of Public Health, as California has the largest Multiracial population of any state.

To maximize subgroup sample sizes, we pooled 2014–2023 data. We categorized Multiracial participants based on all self-identified races, and preserved subgroup (national origin) for monoracial participants. Because BRFSS recommends suppressing results for groups of N<50 participants, we aggregated subgroups of N<50 into broader categories (e.g., Cuban into Hispanic), including multiple subgroup combinations (e.g., Filipino-Mexican into Asian-Hispanic). For residual Multiracial groups where N<50, we followed a previous approach to preserve visibility of groups with known disparities (*8*). We assigned truncated labels in the following order: Native Hawaiian or Pacific Islander [NHPI], American Indian or Alaska Native [AIAN], Black, Asian, and Hispanic, and appended “-Multiple” (e.g., “NHPI-Multiple”). For visualization, we include related subgroups under broader racial and ethnic categories (e.g., Black-White appears in Black, Multiracial, and White categories).

We selected 28 indicators assessed annually to maximize precision. These included lifestyle factors (smoking, alcohol use, physical activity), preventive healthcare (screenings, vaccinations, access to care) chronic conditions (cardiovascular conditions, diabetes, pulmonary conditions, skin cancer, depressive disorder, obesity), self-reported health status (fair/poor general health, frequent poor physical/mental health), and disabilities (vision impairment, cognitive difficulty, mobility limitations). We dichotomized all variables so that higher prevalence indicated poorer health (details in **Supplement**).

We obtained the 2020 American Community Survey 5-year microdata for California adults (18+), and standardized prevalence by age and sex (direct method). We calculated weighted age-sex distributions by biological sex (male/female) and age categories (18-29, 30-44, 45-59, 60+), applied unadjusted prevalence to calculate expected cases, and divided totals by the reference population to obtain standardized prevalence. Using Taylor Series approximation, we calculated relative standard errors (RSE) and dichotomized values (low, <0.3; high ≥0.3) to characterize estimate stability. Within the Multiracial grouping, we calculated crude differences between the highest and lowest prevalence subgroups. For visualizations, we applied min-max normalization to standardized prevalence within each outcome, scaling between 0 and 1. We handled missing data using multiple imputation (20 imputations, 20 iterations). We conducted all analyses in R version 4.4.0 (R Core Team 2024, Vienna, Austria) in 2025. The Stanford University institutional review board exempted this study of deidentified data.

## Results

Among 100,177 participants (50.8% female, 49.2% male; 21.5% ages 18-29 years; 27.1%, 30-44 years; 24.1%, 45-59 years; 27.3%, 60+ years), 36.6% identified as Hispanic (**Table 1**). Among non-Hispanic monoracial groups, 0.6% identified as AIAN; 15.3%, Asian; 5.4%, Black; 0.3%, NHPI; 37.8%, White; and 0.9%, another race; 2.0% identified as non-Hispanic Multiracial.

**Table 1.**
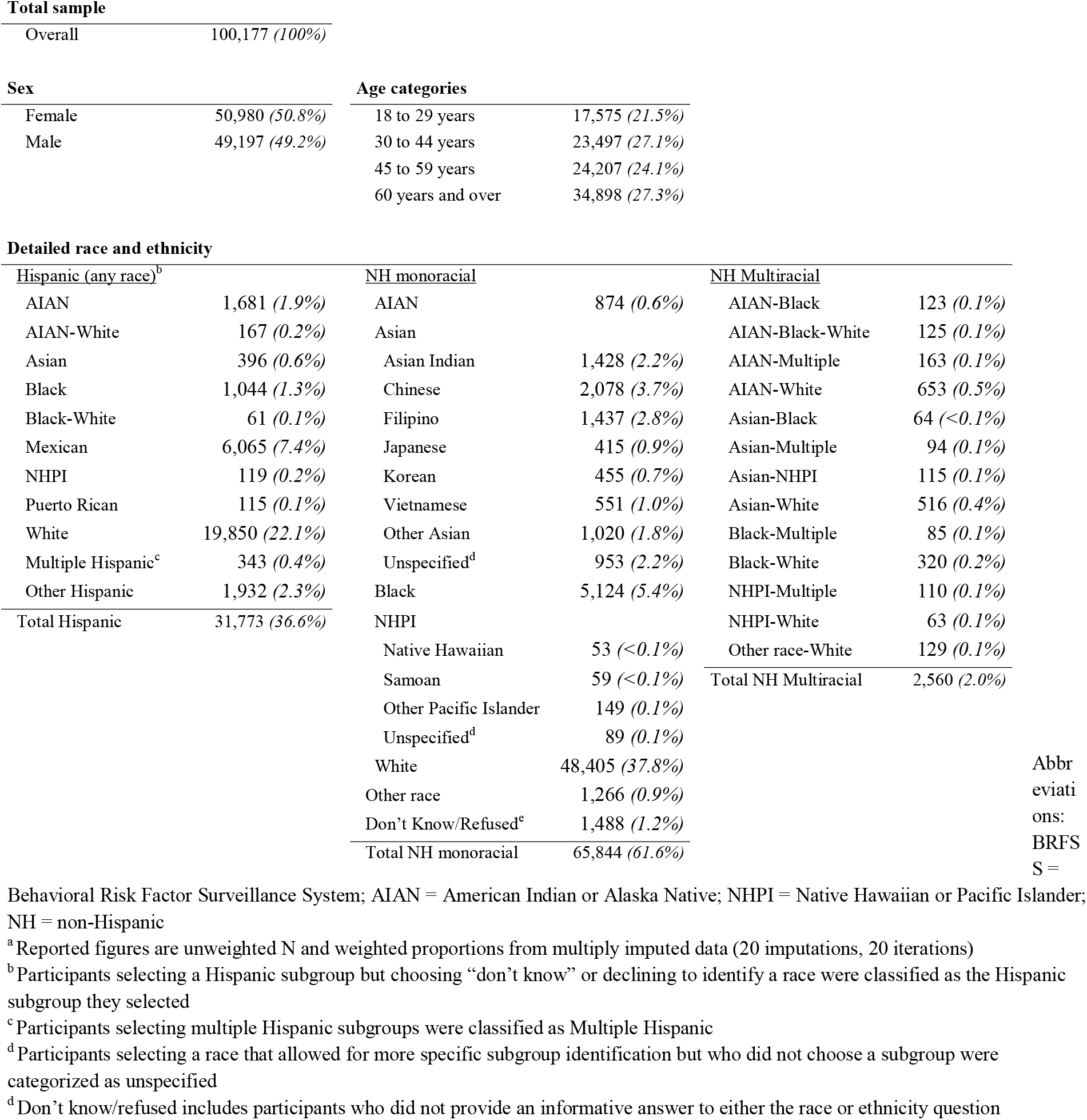
Distribution^a^ of participant sex, age categories, and detailed race and ethnicity, California BRF SS 2014 - 2023.

Notable patterns emerged across the larger racial categories (**Figure 1**). Smoking, chronic conditions, poor general health, and disabilities were highly prevalent among AIAN participants, while binge drinking, no exercise, lack of preventive care, and diabetes were relatively more prevalent among Hispanic participants. Asian participants had high prevalence of never being tested for HIV, but low prevalence of chronic conditions, poor health, and disability. NHPI participants reported low prevalence of poor health and disability, but high prevalence of smoking and binge drinking. Black, Multiracial, and White groupings showed heterogeneity, but disability prevalence was higher among Black adults. **Figure 1** labels only the highest- and lowest-prevalence subgroups for each outcome (all subgroup estimates and 95% CIs available in **Supplement**).

**Fig. 1.**
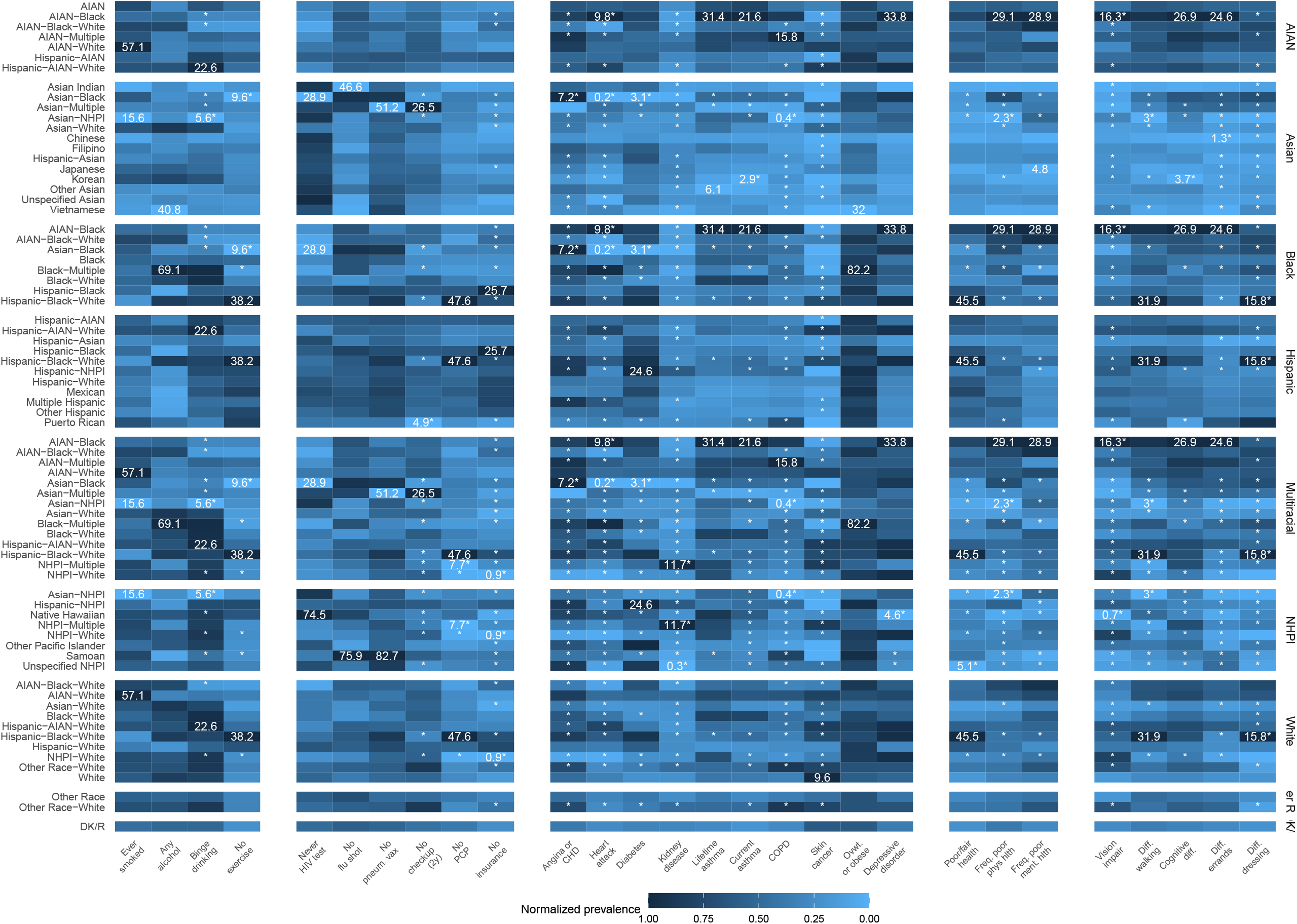

Prevalence varied widely across subgroups. Of 28 outcomes analyzed, Multiracial subgroups had the highest prevalence in 24 (85.7%) and the lowest in 12 (42.9%). AIAN-Black adults had the highest prevalence of nine (32.1%) outcomes – the most of any subgroup – while Hispanic-Black-White adults had the highest in five (17.9%). Asian-Black adults had the lowest prevalence of four outcomes (but the highest for angina or coronary heart disease). Overall, Asian subgroups had the lowest prevalence for 18 (75%) outcomes. Multiracial adults typically had similar or worse health than their monoracial counterparts within each racial grouping.

Within the Multiracial category, other patterns emerged (**Table 2**). Prevalence differences were largest for lifetime smoking (15.6%, Asian-NHPI to 57.1%, AIAN-White), having no primary care provider (7.7%, NHPI-Multiple to 47.6%, Hispanic-Black-White), and self-rated fair or poor health (7.6%, Asian-NHPI to 45.5%, Hispanic-Black-White). Asian Multiracial subgroups (e.g., Asian-NHPI, Asian-Black) most often had the lowest prevalence across lifestyle, chronic, and disability indicators. In contrast, the highest prevalence was typically in Multiracial AIAN, NHPI, and Hispanic subgroups, especially for chronic conditions, access to care, and disability-related measures. Prevalence differences exceeded 20 percentage points for nearly half the outcomes, underscoring the Multiracial population’s heterogeneity.

**Table 2.** Lowest and highest prevalence Multiracial subgroups^a^ and associated prevalence differences by outcome, California BRFSS 2014-2023.

| Health Indicator | Lowest |  | Highest |  | Prevalence difference |
| --- | --- | --- | --- | --- | --- |
|  | Group | Prevalence (95% CI) | Group | Prevalence (95% CI) |  |
| <u>Lifestyle factors</u> |  |  |  |  |  |
| Ever smoked | Asian-NHPI | 15.6 (9.9 - 21.3) | AIAN-White | 57.1 (39.9 - 74.4) | 41.5 |
| Any alcohol use | Asian-Black | 46.9 (41.1 - 52.7) | Black-Multiple | 69.1 (52.2 - 85.9) | 22.1 |
| Binge drinking | Asian-NHPI | 5.6 (2.5 - 8.7) | Hispanic-AIAN-White | 22.6 (8.8 - 36.3) | 16.9 |
| No exercise in last 30 days | Asian-Black | 9.6 (5.1 - 14.1) | Hispanic-Black-White | 38.2 (25.8 - 50.5) | 28.6 |
| <u>Preventive healthcare</u> |  |  |  |  |  |
| Never had HIV test | Asian-Black | 28.9 (23.1 - 34.6) | Asian-NHPI | 69.6 (54.2 - 85.0) | 40.7 |
| No flu vaccine in last 12 months | Black-White | 52.2 (46.8 - 57.6) | Asian-Black | 73.4 (60.2 - 86.7) | 21.2 |
| Never had pneumonia vaccine | Asian-Multiple | 51.2 (46.3 - 56.1) | Asian-Black | 79.1 (65.2 - 93.0) | 27.9 |
| No health insurance | NHPI-White | 0.9 (0.0 - 2.1) | Hispanic-Black-White | 17.9 (5.4 - 30.5) | 17.0 |
| No routine checkup in last 2 years | Asian-Black | 8.1 (3.6 - 12.5) | Asian-Multiple | 26.5 (14.7 - 38.4) | 18.4 |
| No primary care provider | NHPI-Multiple | 7.7 (3.6 - 11.7) | Hispanic-Black-White | 47.6 (33.4 - 61.8) | 39.9 |
| <u>Chronic conditions</u> |  |  |  |  |  |
| Angina or coronary heart disease | Asian-Multiple | 0.0 (0.0 - 0.0) | Asian-Black | 7.2 (0.0 - 16.1) | 7.2 |
| Heart attack | Asian-Black | 0.2 (0.0 - 0.6) | AIAN-Black | 9.8 (0.0 - 19.9) | 9.6 |
| Diabetes | Asian-Black | 3.1 (0.0 - 6.9) | Hispanic-Black-White | 22.7 (10.0 - 35.3) | 19.5 |
| Chronic kidney disease | Asian-Black | 0.5 (0.0 - 1.5) | NHPI-Multiple | 11.7 (3.3 - 20.1) | 11.2 |
| Lifetime asthma | Asian-Multiple | 7.9 (3.1 - 12.7) | AIAN-Black | 31.4 (18.2 - 44.7) | 23.5 |
| Current asthma | Asian-Multiple | 3.9 (0.6 - 7.1) | AIAN-Black | 21.6 (11.2 - 32.1) | 17.8 |
| COPD | Asian-NHPI | 0.4 (0.0 - 1.1) | AIAN-Multiple | 15.8 (7.0 - 24.7) | 15.5 |
| Skin cancer | Asian-Black | 0.0 (0.0 - 0.0) | NHPI-Multiple | 8.1 (0.3 - 15.9) | 8.1 |
| Depressive disorder | NHPI-Multiple | 12.6 (8.0 - 17.3) | AIAN-Black | 33.8 (18.6 - 49.0) | 21.2 |
| Overweight or obese | Asian-NHPI | 51.3 (45.6 - 57.1) | Black-Multiple | 82.2 (69.0 - 95.5) | 30.9 |
| <u>General health status</u> |  |  |  |  |  |
| Fair or poor health status | Asian-NHPI | 7.6 (2.6 - 12.6) | Hispanic-Black-White | 45.5 (32.9 - 58.0) | 37.9 |
| Frequent poor physical health | Asian-NHPI | 2.3 (0.2 - 4.5) | AIAN-Black | 29.1 (16.4 - 41.8) | 26.8 |
| Frequent poor mental health | Black-Multiple | 7.3 (2.5 - 12.0) | AIAN-Black | 28.9 (17.6 - 40.2) | 21.6 |
| <u>Disabilities</u> |  |  |  |  |  |
| Vision impairment | Asian-Multiple | 1.2 (0.0 - 2.7) | AIAN-Black | 16.3 (5.8 - 26.7) | 15.0 |
| Difficulty walking | Asian-NHPI | 3.0 (0.0 - 6.1) | Hispanic-Black-White | 31.9 (16.1 - 47.6) | 28.9 |
| Cognitive difficulty | Asian-White | 9.7 (5.4 - 14.1) | AIAN-Black | 26.9 (15.4 - 38.3) | 17.2 |
| Difficulty running errands | Asian-NHPI | 2.2 (0.0 - 5.0) | AIAN-Black | 24.6 (13.3 - 35.9) | 22.4 |
| Difficulty dressing | NHPI-White | 0.0 (0.0 - 0.0) | Hispanic-Black-White | 15.8 (3.7 - 27.9) | 15.8 |

Abbreviations: BRFSS = Behavioral Risk Factor Surveillance System; AIAN = American Indian or Alaska Native; NHPI = Native Hawaiian or Pacific Islander; HIV = human immunodeficiency virus; COPD = chronic obstructive pulmonary disorder
<sup>a</sup>Sex- and age-group standardized prevalence obtained from multiply imputed data (20 imputations, 20 iterations)

## Discussion

This analysis of a large, state-based survey revealed wide variation in health across racial and ethnic subgroups, including among Multiracial adults. Some Multiracial subgroups (e.g., AIAN-Black, Hispanic-Black-White) had among the highest prevalence of chronic disease, poor general health, and disability across all studied groups, often exceeding rates observed in the monoracial groups they identify with. These findings would be masked by pooling Multiracial participants in a single category or with monoracial groups, as is common practice.

Prior work observed better health among Asian-White adults, a relatively advantaged Multiracial subgroup (*5*). However, we found that other Asian Multiracial subgroups (e.g. Asian-Black, Asian-Multiple, Asian-NHPI) more often had the lowest prevalence of adverse outcomes, calling into question assumptions about which Multiracial subgroups are healthiest. While this study focused on Multiracial participants, variation was also observed across all groupings. These findings underscore the importance of racial and ethnic data in surveillance systems (*6*) and data infrastructure enabling routine disaggregation.

This report had limitations. First, small sample sizes required aggregation of some subgroups, and nearly one-third of prevalence estimates were unstable. BRFSS data also do not allow disaggregation of AIAN, Black, or White subgroups and currently lack a Middle Eastern or North African (MENA) category. Future studies should replicate this analysis using data from more states and after introduction of the MENA category. Second, outcomes were self-reported and may be biased if reporting differs by race or ethnicity. Datasets with objective measures (e.g., physician-reported diagnoses) could help validate findings. Finally, BRFSS response rates declined in 2020-2021 due to the COVID-19 pandemic, and prior analyses have shown racial and ethnic response rate differences in BRFSS (*9*). We did not assess the impact of these potential selection biases.

Disaggregated race and ethnicity data are valuable for identifying at-risk subgroups, and prevention pograms may be most effective if designed to support subgroups with highest burden (*10*). Improving data infrastructure to support subgroup-level reporting is essential for effective disease prevention programs.

## Data Availability

Original data used for this analysis are available upon request from the California Department of Public Health. Code used for this analysis is available online at https://github.com/lamhine/brfss_disagg.

## Acknowledgements

The authors would like to thank Vanessa Miguelino-Keasling and Nicholas Cuvelier for their assistance in accessing the data for this analysis

## References

1. US Census Bureau. Census.gov. 2021 [cited 2022 Mar 7]. Improved Race and Ethnicity Measures Reveal U.S. Population Is Much More Multiracial. Available from: https://www.census.gov/library/stories/2021/08/improved-race-ethnicity-measures-reveal-united-states-population-much-more-multiracial.html

2. Giano Z, Wheeler DL, Hubach RD. The frequencies and disparities of adverse childhood experiences in the U.S. BMC Public Health. 2020 Dec;20(1):1327.

3. National Center for Health Statistics. Summary Health Statistics: National Health Interview Survey [Internet]. Centers for Disease Control and Prevention; 2018 [cited 2024 Nov 21]. Available from: https://wwwn.cdc.gov/NHISDataQueryTool/SHS_adult/index.html

4. Town M, Eke P, Zhao G, Thomas CW, Hsia J, Pierannunzi C, et al. Racial and Ethnic Differences in Social Determinants of Health and Health-Related Social Needs Among Adults — Behavioral Risk Factor Surveillance System, United States, 2022. MMWR Morb Mortal Wkly Rep. 2024 Mar 7;73(9):204–8.

5. Bratter JL, Gorman BK. Does Multiracial Matter? A Study of Racial Disparities in Self-Rated Health. Demography. 2011 Feb;48(1):127–52.

6. Ponce NA, Becker T, Shimkhada R. Breaking Barriers with Data Equity: The Essential Role of Data Disaggregation in Achieving Health Equity. Annu Rev Public Health [Internet]. 2025 Jan 30 [cited 2025 Feb 3]; Available from: https://www.annualreviews.org/content/journals/10.1146/annurev-publhealth-072523-093838

7. Lam□Hine T, Forthal S, Johnson CY, Chin HB. Asking MultiCrit Questions: A Reflexive and Critical Framework to Promote Health Data Equity for the Multiracial Population. Milbank Q. 2024 Jun;102(2):398–428.

8. Lam-Hine T, Thakur N, Saperstein A, Mujahid MS, Rehkopf DH. Asthma Prevalence in the Disaggregated Multiracial Population of California. JAMA Netw Open. 2024 Dec 9;7(12):e2449588.

9. Schneider KL, Clark MA, Rakowski W, Lapane KL. Evaluating the impact of non-response bias in the Behavioral Risk Factor Surveillance System (BRFSS). J Epidemiol Community Health. 2012 Apr;66(4):290–5.

10. Bauer UE, Briss PA, Goodman RA, Bowman BA. Prevention of chronic disease in the 21st century: elimination of the leading preventable causes of premature death and disability in the USA. The Lancet. 2014 Jul;384(9937):45–52.

