## Supplement for "Chronic Disease and Risk Factor Prevalence in Multiracial Subgroups: California, 2014–2023"

**Appendix A.** Operationalization of health indicators by category, California BRFSS 2014-2023

| **Lifestyle factors**   - Ever smoked: current or former calculated smoking status - No exercise in last 30 days: “no” response to “*During the past month, other than your regular job, did you participate in any physical activities or exercises?*” - Any alcohol use: had at least one drink of alcohol in past 30 days - Binge drinking: calculated sex-specific binge drinking status   **Preventive healthcare**   - Never had HIV test: “no” response to *“Have you ever been tested for HIV?”* - No flu vaccine in last 12 months: “no” response to *“During the past 12 months, have you had either [nasal or injected] flu vaccine?”* - Never had pneumonia vaccine: “no” response to *“Have you ever had a pneumonia shot also known as a pneumococcal vaccine?”* - No health insurance: “no” response to *“Do you have any kind of health care coverage, including health insurance, prepaid plans such as HMOs, or government plans such as Medicare, or Indian Health Service?”* - No routine checkup in last 2 years: “2 years” or more” response to *“About how long has it been since you last visited a doctor for a routine checkup?”* - No primary care provider: “no” response to *“Do you have one person you think of as your personal doctor or health care provider?”*   **Chronic conditions** (*“Has a doctor, nurse, or other health professional ever told you that you had…”*)   - Angina or coronary heart disease: “yes” response to *“… angina or coronary heart disease?”* - Heart attack: “yes” response to *“… a heart attack, also called a myocardial infarction?”* - Diabetes: “yes” response to *“… diabetes?”*, not including gestational, borderline, or pre-diabetes - Chronic kidney disease: “yes” response to *“… kidney disease, not including kidney stones, bladder infection, or incontinence?”* - Lifetime asthma: “yes” response to *“… asthma?”* | - Current asthma: “yes” response to *“… currently have asthma?”* - Chronic obstructive pulmonary disorder (COPD): “yes” response to *“… chronic obstructive pulmonary disease, COPD, emphysema, or chronic bronchitis?”* - Skin cancer: “yes” response to *“… skin cancer?”* - Depressive disorder: “yes” response to *“… a depressive disorder (including depression, major depression, dysthymia, or minor depression?”* - Overweight or obese: body mass index > 25.0   **General health status**   - Fair or poor health status: “fair” or “poor” response to *“Would you say that in general your health is…”* - Frequent poor physical health: “15 or more” response to *“Now thinking about your physical health, which includes physical illness and injury, for how many days during the past 30 days was your physical health not good?”* - Frequent poor mental health: “15 or more” response to *“Now thinking about your mental health, which includes stress, depression, and problems with emotions, for how many days during the past 30 days was your mental health not good?”*   **Disabilities**   - Vision impairment: “yes” response to: *“Are you blind or do you have serious difficulty seeing, even when wearing glasses?”* - Difficulty walking: “yes” response to: *“Do you have serious difficulty walking or climbing stairs?”* - Cognitive difficulty: “yes” response to: *“Because of a physical, mental, or emotional condition, do you have serious difficulty concentrating, remembering, or making decisions?”* - Difficulty running errands: “yes” response to: *“Because of a physical, mental, or emotional condition, do you have difficulty doing errands alone such as visiting a doctor’s office or shopping?”* - Difficulty dressing: “yes” response to: *“Do you have difficulty dressing or bathing?”* |
| --- | --- |

| **Appendix B.** Distribution^a^ of health indicators in study population, overall and stratified by sex and by age group, California BRFSS 2014-2023 | | | | | | | | | | | | | | | | | | | | | | | | | | | | |
| --- | --- | --- | --- | --- | --- | --- | --- | --- | --- | --- | --- | --- | --- | --- | --- | --- | --- | --- | --- | --- | --- | --- | --- | --- | --- | --- | --- | --- |
|  | Angina or CHD | Lifetime asthma | Current asthma | Skin cancer | No checkup (2y) | Kidney disease | COPD | Cognitive diff. | Diff. dressing | Diff. errands | Diff. walking | Diabetes | Any alcohol | Binge drinking | No exercise | Poor/fair health | Never HIV test | No insurance | No flu shot | Depressive disorder | Heart attack | Freq. poor ment. hlth | Freq. poor phys hlth | Ovwt. or obese | No PCP | No pneum. vax | Ever smoked | Vision impair |
| AIAN | 4.6 (2.8–6.4) | 23.4 (19.2–27.6) | 17.3 (13.5–21.2) | 3.1 (1.5–4.6) | 13.3 (10.0–16.5) | 3.1 (1.7–4.4) | 7.0 (4.7–9.2) | 17.4 (13.7–21.2) | 8.4 (5.9–11.0) | 13.7 (10.5–17.0) | 21.3 (17.3–25.2) | 16.5 (12.6–20.4) | 48.2 (43.4–53.1) | 16.7 (12.9–20.5) | 22.1 (17.4–26.8) | 26.6 (21.9–31.3) | 43.3 (38.5–48.1) | 5.3 (3.1–7.5) | 61.8 (56.8–66.8) | 19.7 (16.0–23.5) | 5.7 (3.8–7.6) | 17.1 (13.2–21.0) | 18.1 (14.0–22.3) | 70.8 (66.5–75.2) | 17.0 (13.4–20.6) | 59.8 (55.1–64.6) | 45.6 (40.6–50.6) | 7.2 (4.9–9.5) |
| AIAN-Black | 5.9 (1.3–10.4) | 31.4 (20.7–42.2) | 21.6 (12.5–30.7) | 0.8 (0.0–2.3) | 16.0 (8.2–23.7) | 1.6 (0.0–3.5) | 13.6 (7.3–19.9) | 26.9 (15.4–38.3) | 9.3 (3.4–15.1) | 24.6 (13.3–35.9) | 29.6 (20.2–38.9) | 19.6 (10.8–28.3) | 55.2 (46.8–63.6) | 10.3 (2.4–18.1) | 17.7 (10.3–25.1) | 35.7 (23.1–48.2) | 38.3 (29.1–47.5) | 9.7 (1.1–18.2) | 65.9 (55.8–76.0) | 33.8 (23.0–44.6) | 9.8 (2.7–16.9) | 28.9 (18.2–39.6) | 29.1 (17.8–40.4) | 64.5 (53.6–75.5) | 18.5 (10.3–26.8) | 61.8 (51.6–72.1) | 34.7 (23.8–45.6) | 16.3 (6.1–26.4) |
| AIAN-Black-White | 2.5 (0.3–4.8) | 17.8 (9.8–25.8) | 12.1 (5.7–18.5) | 3.2 (0.0–7.7) | 15.4 (8.2–22.6) | 1.9 (0.0–4.4) | 5.1 (1.4–8.8) | 19.2 (11.2–27.2) | 11.7 (5.3–18.1) | 14.0 (7.2–20.9) | 21.6 (13.6–29.6) | 17.5 (7.5–27.4) | 61.7 (50.2–73.2) | 8.1 (2.9–13.3) | 13.4 (7.5–19.2) | 23.0 (13.3–32.7) | 32.1 (21.9–42.3) | 12.5 (4.8–20.1) | 63.3 (51.5–75.1) | 22.0 (14.2–29.9) | 1.0 (0.0–2.6) | 27.2 (15.9–38.4) | 22.3 (12.3–32.3) | 76.2 (65.9–86.5) | 12.3 (6.7–18.0) | 68.3 (58.2–78.5) | 46.5 (34.9–58.2) | 6.1 (1.9–10.3) |
| AIAN-Multiple | 6.6 (1.9–11.3) | 24.5 (17.2–31.8) | 16.8 (10.7–23.0) | 4.0 (0.5–7.4) | 17.8 (10.2–25.5) | 4.6 (0.6–8.6) | 15.8 (8.8–22.9) | 23.8 (16.0–31.7) | 10.5 (4.0–17.1) | 20.4 (12.4–28.4) | 20.2 (12.2–28.1) | 13.4 (6.5–20.3) | 50.4 (40.3–60.5) | 14.6 (7.0–22.2) | 22.8 (13.7–31.9) | 29.4 (19.9–38.9) | 38.9 (29.1–48.7) | 8.4 (3.6–13.3) | 64.0 (54.5–73.6) | 22.4 (14.1–30.6) | 5.7 (1.5–9.9) | 9.8 (5.0–14.6) | 17.5 (10.7–24.2) | 63.3 (53.9–72.8) | 18.9 (11.4–26.4) | 62.5 (53.4–71.5) | 42.4 (33.7–51.1) | 9.4 (3.7–15.1) |
| AIAN-White | 6.0 (3.6–8.5) | 24.7 (19.5–29.9) | 14.6 (10.4–18.8) | 7.7 (5.0–10.3) | 18.1 (13.1–23.1) | 5.8 (3.1–8.6) | 10.4 (7.3–13.4) | 21.1 (16.7–25.4) | 10.1 (6.9–13.3) | 14.8 (11.0–18.6) | 22.1 (17.4–26.7) | 14.0 (9.9–18.0) | 49.6 (43.8–55.4) | 12.0 (8.9–15.0) | 19.0 (14.5–23.6) | 24.6 (19.4–29.8) | 46.5 (40.8–52.3) | 4.6 (2.0–7.1) | 60.8 (55.4–66.2) | 26.4 (21.8–31.1) | 4.9 (3.0–6.8) | 22.4 (17.7–27.1) | 18.4 (14.1–22.6) | 67.2 (61.5–73.0) | 17.8 (13.3–22.3) | 63.8 (58.9–68.7) | 57.1 (51.4–62.8) | 4.8 (3.0–6.7) |
| Asian Indian | 3.8 (2.0–5.6) | 9.5 (6.8–12.2) | 6.2 (3.8–8.7) | 0.4 (0.0–1.0) | 10.2 (7.5–12.9) | 1.7 (0.7–2.7) | 2.0 (0.9–3.0) | 4.5 (3.0–6.1) | 1.5 (0.4–2.7) | 2.7 (1.4–4.1) | 5.0 (3.1–6.9) | 13.6 (10.7–16.6) | 47.8 (43.5–52.0) | 6.6 (5.1–8.1) | 14.9 (11.8–17.9) | 7.5 (5.3–9.7) | 69.0 (65.0–72.9) | 3.8 (1.9–5.6) | 46.6 (42.3–50.9) | 5.0 (3.3–6.6) | 3.6 (1.9–5.4) | 5.7 (3.8–7.6) | 2.8 (1.6–4.0) | 48.3 (44.0–52.7) | 10.7 (8.8–12.6) | 67.3 (63.0–71.6) | 16.6 (13.7–19.6) | 0.8 (0.3–1.4) |
| Asian-Black | 7.2 (0.0–16.1) | 18.5 (5.2–31.7) | 11.9 (1.4–22.3) | 0.0 (0.0–0.0) | 8.1 (1.8–14.4) | 0.5 (0.0–1.5) | 7.4 (0.0–16.2) | 18.4 (8.2–28.7) | 11.0 (1.7–20.3) | 15.2 (6.1–24.3) | 15.5 (2.0–29.0) | 3.1 (0.0–7.6) | 46.9 (30.1–63.8) | 10.9 (2.1–19.7) | 9.6 (1.7–17.wat5) | 12.0 (1.2–22.8) | 28.9 (14.8–42.9) | 8.8 (0.0–18.9) | 73.4 (61.7–85.1) | 29.8 (15.7–43.9) | 0.2 (0.0–0.6) | 12.8 (3.1–22.5) | 21.3 (8.6–34.0) | 67.4 (54.1–80.7) | 21.9 (12.4–31.3) | 79.1 (69.6–88.5) | 40.7 (23.5–58.0) | 2.0 (0.1–3.9) |
| Asian-Multiple | 0.0 (0.0–0.0) | 7.9 (2.8–13.0) | 3.9 (0.6–7.1) | 5.2 (0.0–12.2) | 26.5 (14.7–38.4) | 4.2 (0.0–9.3) | 1.5 (0.0–3.5) | 15.2 (5.8–24.5) | 6.9 (0.5–13.4) | 13.3 (4.8–21.9) | 11.4 (3.2–19.5) | 10.2 (1.9–18.4) | 49.2 (35.7–62.7) | 11.9 (4.5–19.4) | 17.4 (9.7–25.0) | 11.2 (3.3–19.2) | 62.4 (48.3–76.5) | 3.9 (0.0–8.4) | 69.1 (57.1–81.0) | 17.9 (8.8–27.0) | 4.4 (0.0–10.4) | 14.0 (5.9–22.2) | 8.0 (1.5–14.5) | 65.6 (54.1–77.1) | 20.6 (11.1–30.1) | 51.2 (37.3–65.1) | 27.9 (16.8–39.1) | 1.2 (0.0–2.7) |
| Asian-NHPI | 3.0 (0.0–6.1) | 19.1 (9.7–28.4) | 12.1 (3.6–20.5) | 0.5 (0.0–1.5) | 9.6 (3.0–16.2) | 2.2 (0.0–5.2) | 0.4 (0.0–1.1) | 11.2 (2.3–20.2) | 0.2 (0.0–0.5) | 2.2 (0.0–5.0) | 3.0 (0.0–6.1) | 7.0 (0.9–13.2) | 47.2 (36.5–57.8) | 5.6 (0.8–10.4) | 15.8 (7.2–24.5) | 7.6 (2.2–13.0) | 69.6 (58.8–80.5) | 6.8 (0.5–13.1) | 53.0 (41.7–64.3) | 21.4 (9.8–33.0) | 3.5 (0.0–7.4) | 17.0 (6.6–27.5) | 2.3 (0.2–4.5) | 51.3 (39.6–63.0) | 19.7 (10.8–28.6) | 70.3 (58.9–81.7) | 15.6 (7.1–24.2) | 5.5 (0.0–11.9) |
| Asian-White | 2.7 (0.0–6.5) | 17.0 (12.2–21.8) | 7.0 (3.7–10.3) | 4.9 (0.9–9.0) | 16.7 (12.3–21.2) | 2.7 (0.1–5.3) | 3.5 (0.0–7.3) | 9.7 (5.0–14.5) | 3.0 (0.0–6.8) | 4.5 (0.4–8.5) | 7.2 (2.4–12.0) | 7.1 (3.4–10.9) | 65.4 (58.7–72.2) | 18.3 (13.4–23.2) | 13.5 (8.1–18.9) | 10.0 (5.0–15.1) | 46.6 (39.7–53.6) | 2.4 (1.0–3.8) | 53.1 (46.1–60.1) | 19.1 (13.0–25.3) | 1.9 (0.0–4.2) | 11.6 (6.8–16.3) | 5.5 (1.4–9.6) | 54.4 (47.2–61.6) | 17.5 (13.4–21.5) | 67.1 (61.1–73.1) | 42.4 (35.0–49.8) | 2.7 (0.0–6.4) |
| Black | 3.1 (2.5–3.7) | 20.9 (19.3–22.6) | 13.1 (11.8–14.5) | 0.6 (0.3–0.9) | 10.3 (9.0–11.5) | 3.1 (2.5–3.8) | 5.5 (4.7–6.3) | 13.6 (12.3–14.9) | 6.6 (5.7–7.5) | 9.7 (8.5–10.8) | 17.7 (16.3–19.1) | 14.0 (12.7–15.4) | 54.0 (52.1–56.0) | 14.9 (13.5–16.2) | 23.7 (22.0–25.4) | 19.0 (17.5–20.4) | 40.9 (39.0–42.8) | 6.5 (5.4–7.5) | 66.2 (64.4–68.0) | 17.1 (15.6–18.5) | 3.1 (2.5–3.7) | 14.1 (12.8–15.4) | 13.0 (11.7–14.2) | 73.9 (72.3–75.6) | 16.5 (15.1–17.8) | 69.1 (67.4–70.7) | 37.8 (35.8–39.7) | 6.9 (5.8–8.0) |
| Black-Multiple | 5.8 (0.0–13.5) | 15.4 (7.5–23.2) | 6.7 (0.1–13.3) | 0.3 (0.0–1.0) | 9.1 (1.6–16.6) | 1.3 (0.0–3.2) | 7.9 (0.1–15.7) | 10.4 (2.3–18.6) | 10.4 (1.9–19.0) | 12.9 (5.0–20.8) | 21.1 (10.7–31.5) | 12.3 (3.1–21.5) | 69.1 (57.6–80.6) | 21.9 (10.1–33.8) | 12.8 (2.6–23.0) | 15.9 (5.0–26.9) | 33.2 (18.2–48.1) | 5.2 (0.9–9.5) | 66.2 (53.0–79.3) | 22.0 (10.3–33.8) | 9.4 (0.0–19.5) | 7.3 (2.3–12.2) | 17.8 (7.3–28.2) | 82.2 (73.2–91.3) | 13.6 (6.2–21.1) | 60.0 (47.2–72.8) | 33.1 (20.3–46.0) | 7.2 (0.0–15.3) |
| Black-White | 4.5 (1.0–8.1) | 26.5 (18.9–34.1) | 17.3 (10.5–24.0) | 3.6 (0.0–8.3) | 17.9 (11.2–24.7) | 2.6 (0.2–4.9) | 7.2 (2.5–11.9) | 14.9 (9.0–20.9) | 6.2 (1.9–10.5) | 9.1 (5.0–13.1) | 13.2 (7.5–19.0) | 7.4 (2.4–12.4) | 61.6 (53.6–69.6) | 21.6 (14.9–28.4) | 18.4 (11.1–25.7) | 14.7 (8.8–20.6) | 46.2 (37.8–54.6) | 6.6 (2.9–10.4) | 52.2 (43.3–61.1) | 21.1 (13.6–28.5) | 4.1 (0.7–7.6) | 19.6 (12.9–26.2) | 10.5 (5.2–15.8) | 69.6 (62.0–77.3) | 20.9 (14.0–27.8) | 61.9 (53.7–70.1) | 36.7 (28.8–44.6) | 4.0 (1.3–6.6) |
| Chinese | 1.5 (0.7–2.3) | 9.5 (7.4–11.5) | 5.0 (3.3–6.8) | 0.1 (0.0–0.3) | 11.9 (10.1–13.8) | 1.9 (0.9–3.0) | 2.1 (1.0–3.2) | 4.5 (3.0–5.9) | 0.5 (0.1–0.9) | 1.3 (0.5–2.1) | 4.7 (3.1–6.2) | 7.9 (6.0–9.8) | 47.5 (44.5–50.6) | 7.8 (6.3–9.2) | 15.4 (13.0–17.8) | 7.4 (5.6–9.1) | 68.8 (66.0–71.7) | 3.1 (2.1–4.1) | 53.6 (50.4–56.8) | 6.3 (4.7–8.0) | 1.6 (0.7–2.6) | 4.9 (3.6–6.1) | 4.7 (3.3–6.1) | 39.0 (35.9–42.1) | 13.8 (12.0–15.6) | 69.8 (67.1–72.5) | 18.6 (16.0–21.2) | 2.4 (1.2–3.5) |
| DK/R | 2.8 (1.8–3.7) | 12.7 (10.5–14.9) | 6.6 (5.0–8.1) | 5.9 (4.2–7.5) | 14.4 (12.1–16.7) | 3.7 (2.4–5.0) | 4.5 (3.2–5.9) | 11.0 (8.4–13.6) | 3.4 (2.1–4.7) | 7.4 (5.4–9.3) | 13.2 (10.7–15.7) | 9.8 (7.4–12.3) | 55.2 (51.5–59.0) | 14.4 (11.8–17.0) | 15.8 (13.3–18.4) | 14.0 (11.6–16.3) | 51.5 (47.9–55.0) | 6.9 (5.1–8.7) | 63.5 (59.9–67.0) | 15.1 (12.2–17.9) | 2.7 (1.7–3.6) | 11.2 (9.2–13.1) | 12.6 (9.9–15.3) | 63.2 (59.5–66.9) | 23.2 (20.1–26.3) | 68.0 (64.7–71.2) | 34.7 (31.3–38.2) | 4.7 (3.2–6.2) |
| Filipino | 4.4 (2.9–5.9) | 16.8 (14.0–19.5) | 9.1 (6.9–11.2) | 0.5 (0.0–1.0) | 11.2 (8.9–13.4) | 3.5 (2.2–4.8) | 2.6 (1.2–4.0) | 7.4 (5.7–9.1) | 3.1 (1.8–4.3) | 4.4 (3.1–5.8) | 10.3 (8.1–12.6) | 15.6 (13.2–18.1) | 47.4 (43.9–50.9) | 12.2 (10.0–14.4) | 19.2 (16.5–22.0) | 14.3 (11.6–16.9) | 65.2 (61.9–68.5) | 5.9 (4.3–7.4) | 49.5 (46.1–53.0) | 8.4 (6.5–10.2) | 4.5 (2.8–6.1) | 9.4 (7.4–11.3) | 8.5 (6.4–10.6) | 57.3 (53.9–60.7) | 14.2 (12.0–16.5) | 65.5 (62.3–68.6) | 30.7 (27.5–33.9) | 4.3 (2.6–5.9) |
| Hispanic-AIAN | 3.7 (2.5–5.0) | 16.2 (13.6–18.8) | 10.0 (7.9–12.1) | 0.6 (0.2–1.1) | 17.4 (14.8–19.9) | 3.5 (2.2–4.8) | 4.8 (3.3–6.2) | 14.4 (12.1–16.7) | 6.3 (4.5–8.1) | 9.4 (7.3–11.5) | 18.3 (15.7–20.8) | 15.4 (13.1–17.6) | 49.5 (46.2–52.8) | 18.8 (16.4–21.3) | 27.4 (24.4–30.4) | 31.5 (28.3–34.7) | 54.2 (51.1–57.4) | 16.1 (14.0–18.3) | 64.6 (61.4–67.8) | 17.3 (14.8–19.9) | 4.8 (3.5–6.0) | 14.6 (12.2–17.1) | 15.3 (12.8–17.8) | 75.9 (73.0–78.7) | 29.9 (27.1–32.8) | 71.8 (68.9–74.6) | 38.3 (35.1–41.5) | 8.1 (6.1–10.0) |
| Hispanic-AIAN-White | 4.3 (1.1–7.5) | 17.9 (10.5–25.2) | 10.2 (4.9–15.4) | 8.0 (2.4–13.7) | 14.4 (7.3–21.6) | 2.3 (0.0–4.8) | 6.8 (2.3–11.2) | 20.4 (13.5–27.4) | 7.8 (3.0–12.6) | 15.3 (8.0–22.7) | 21.5 (12.4–30.6) | 11.2 (6.1–16.4) | 59.5 (48.8–70.3) | 22.6 (13.2–31.9) | 22.7 (15.1–30.2) | 27.6 (18.5–36.7) | 41.4 (31.2–51.5) | 8.4 (4.0–12.9) | 63.1 (52.7–73.5) | 32.0 (23.0–41.1) | 7.6 (2.9–12.3) | 17.6 (10.2–24.9) | 15.2 (8.9–21.5) | 72.9 (64.6–81.2) | 18.9 (11.2–26.7) | 67.9 (60.1–75.7) | 43.2 (32.2–54.3) | 10.6 (3.7–17.4) |
| Hispanic-Asian | 3.6 (0.8–6.4) | 11.3 (7.5–15.2) | 6.6 (3.8–9.3) | 1.1 (0.0–2.3) | 13.8 (9.3–18.3) | 3.5 (0.7–6.2) | 3.4 (1.1–5.8) | 9.3 (5.1–13.5) | 1.3 (0.0–3.0) | 2.9 (0.9–5.0) | 8.6 (4.5–12.6) | 17.2 (11.6–22.8) | 50.4 (43.9–56.9) | 13.3 (8.9–17.8) | 16.3 (11.2–21.4) | 14.8 (10.0–19.5) | 61.9 (55.6–68.2) | 9.5 (5.3–13.8) | 57.0 (50.3–63.8) | 9.4 (6.0–12.8) | 3.1 (0.3–5.9) | 11.3 (7.2–15.5) | 8.5 (4.9–12.1) | 53.9 (47.2–60.5) | 18.8 (14.1–23.6) | 68.2 (62.2–74.2) | 27.1 (21.4–32.8) | 3.9 (1.0–6.7) |
| Hispanic-Black | 3.3 (1.9–4.6) | 14.5 (11.4–17.6) | 9.5 (6.9–12.1) | 1.9 (0.5–3.4) | 18.2 (15.2–21.1) | 3.3 (1.8–4.7) | 4.2 (2.4–6.0) | 13.7 (10.9–16.5) | 6.4 (4.1–8.7) | 7.5 (5.2–9.9) | 18.3 (14.9–21.8) | 19.0 (15.7–22.4) | 42.5 (38.3–46.6) | 15.6 (12.9–18.3) | 25.6 (22.0–29.1) | 31.7 (27.7–35.6) | 56.2 (52.1–60.3) | 25.7 (22.3–29.2) | 70.0 (66.1–73.8) | 14.2 (11.3–17.1) | 3.4 (1.9–5.0) | 10.8 (8.3–13.4) | 14.3 (11.4–17.2) | 76.7 (73.5–79.9) | 32.8 (29.3–36.3) | 73.3 (69.8–76.8) | 35.8 (31.8–39.7) | 7.2 (5.1–9.3) |
| Hispanic-Black-White | 6.0 (0.0–13.5) | 16.0 (5.3–26.7) | 11.7 (2.8–20.5) | 7.0 (0.0–14.8) | 11.2 (3.7–18.8) | 3.2 (0.0–8.5) | 5.4 (0.0–11.3) | 19.7 (9.0–30.3) | 15.8 (3.7–27.9) | 7.1 (0.0–14.8) | 31.9 (16.1–47.6) | 22.7 (10.0–35.3) | 64.6 (52.6–76.5) | 18.6 (8.0–29.1) | 38.2 (30.3–46.0) | 45.5 (33.7–57.3) | 49.0 (34.4–63.7) | 17.9 (5.4–30.5) | 64.7 (51.4–77.9) | 32.5 (17.3–47.7) | 6.0 (0.0–13.5) | 15.8 (4.8–26.8) | 13.4 (3.0–23.8) | 68.6 (57.9–79.4) | 47.6 (33.4–61.8) | 78.5 (67.7–89.3) | 23.2 (12.9–33.5) | 9.0 (1.4–16.5) |
| Hispanic-NHPI | 6.1 (0.9–11.4) | 15.7 (7.9–23.5) | 11.4 (4.6–18.1) | 0.0 (0.0–0.0) | 18.0 (10.3–25.7) | 7.2 (1.3–13.1) | 7.9 (1.5–14.2) | 10.5 (3.6–17.4) | 3.7 (0.0–7.6) | 7.6 (0.2–15.1) | 14.3 (7.1–21.4) | 24.6 (15.2–34.0) | 45.8 (35.2–56.5) | 17.7 (9.5–26.0) | 30.3 (19.4–41.2) | 19.2 (10.9–27.5) | 52.0 (42.2–61.8) | 15.2 (6.6–23.9) | 63.9 (52.8–75.0) | 16.6 (7.7–25.4) | 2.8 (0.0–6.6) | 8.0 (3.1–12.9) | 16.8 (8.1–25.5) | 79.7 (71.2–88.1) | 25.4 (15.5–35.3) | 72.5 (62.4–82.5) | 35.3 (24.8–45.7) | 9.9 (2.1–17.8) |
| Hispanic-White | 3.0 (2.6–3.4) | 10.8 (10.2–11.4) | 6.5 (6.0–7.0) | 2.3 (2.0–2.6) | 16.6 (15.9–17.3) | 3.1 (2.8–3.5) | 4.2 (3.7–4.6) | 11.4 (10.8–12.1) | 5.0 (4.6–5.5) | 6.9 (6.4–7.5) | 15.1 (14.4–15.8) | 15.8 (15.0–16.5) | 46.2 (45.3–47.2) | 14.7 (14.0–15.4) | 28.4 (27.5–29.3) | 30.4 (29.5–31.4) | 58.3 (57.3–59.2) | 19.3 (18.5–20.0) | 63.2 (62.2–64.1) | 13.1 (12.4–13.7) | 3.6 (3.2–4.0) | 10.5 (9.8–11.1) | 13.6 (12.9–14.3) | 74.6 (73.8–75.4) | 28.2 (27.4–29.0) | 74.1 (73.2–74.9) | 30.3 (29.4–31.2) | 6.7 (6.2–7.2) |
| Japanese | 2.5 (0.8–4.1) | 10.6 (6.8–14.4) | 5.4 (2.7–8.1) | 2.6 (0.1–5.2) | 15.7 (10.8–20.6) | 3.9 (0.8–7.0) | 2.5 (0.7–4.4) | 5.9 (2.8–9.0) | 1.9 (0.1–3.7) | 4.5 (1.3–7.8) | 5.0 (2.8–7.3) | 11.9 (8.2–15.6) | 56.9 (50.8–62.9) | 13.7 (9.1–18.3) | 16.6 (12.2–21.0) | 7.2 (4.2–10.2) | 60.6 (54.1–67.0) | 4.1 (1.4–6.8) | 53.0 (46.5–59.5) | 9.6 (5.6–13.7) | 1.6 (0.1–3.0) | 4.8 (2.4–7.2) | 6.2 (3.1–9.3) | 54.2 (48.0–60.4) | 14.0 (9.5–18.5) | 67.3 (62.0–72.6) | 37.3 (30.7–43.9) | 3.9 (1.4–6.5) |
| Korean | 1.3 (0.0–2.6) | 8.2 (4.2–12.3) | 2.9 (0.3–5.5) | 0.0 (0.0–0.0) | 16.8 (12.0–21.5) | 3.4 (0.3–6.6) | 3.4 (0.1–6.7) | 3.7 (0.9–6.4) | 2.5 (0.0–5.0) | 3.5 (0.7–6.3) | 5.8 (1.8–9.9) | 11.3 (6.5–16.2) | 62.0 (54.4–69.6) | 16.7 (11.5–22.0) | 17.4 (11.8–23.0) | 11.5 (6.7–16.4) | 65.3 (58.5–72.2) | 6.5 (3.4–9.6) | 51.8 (44.0–59.6) | 7.6 (4.2–11.0) | 0.8 (0.1–1.5) | 6.3 (3.1–9.5) | 3.6 (1.4–5.8) | 40.4 (33.1–47.6) | 19.2 (14.6–23.8) | 68.9 (61.7–76.1) | 42.9 (35.6–50.3) | 4.5 (0.7–8.3) |
| Mexican | 2.2 (1.7–2.7) | 9.6 (8.6–10.6) | 5.6 (4.8–6.4) | 0.8 (0.5–1.1) | 15.8 (14.7–17.0) | 3.3 (2.7–4.0) | 3.1 (2.4–3.7) | 13.5 (12.3–14.7) | 5.4 (4.6–6.2) | 7.9 (6.9–8.8) | 16.3 (15.1–17.6) | 19.0 (17.7–20.4) | 42.4 (40.7–44.1) | 16.4 (15.1–17.6) | 32.7 (31.0–34.4) | 35.2 (33.6–36.9) | 61.7 (60.0–63.4) | 19.5 (18.2–20.8) | 65.4 (63.8–67.1) | 12.8 (11.7–14.0) | 3.5 (2.8–4.1) | 11.1 (9.9–12.3) | 13.3 (12.1–14.5) | 78.9 (77.5–80.2) | 31.5 (30.0–33.1) | 76.3 (74.8–77.8) | 28.9 (27.3–30.4) | 9.1 (8.1–10.2) |
| Multiple Hispanic | 6.3 (1.3–11.3) | 10.4 (6.4–14.4) | 6.9 (3.5–10.3) | 1.6 (0.1–3.1) | 16.3 (11.0–21.5) | 1.8 (0.0–3.6) | 4.3 (1.8–6.8) | 13.7 (8.6–18.9) | 6.3 (3.0–9.6) | 8.9 (4.4–13.3) | 13.8 (9.2–18.3) | 13.7 (8.5–19.0) | 43.0 (35.4–50.6) | 13.2 (9.0–17.4) | 23.5 (16.9–30.1) | 24.5 (17.7–31.3) | 58.3 (50.6–66.0) | 20.0 (13.9–26.1) | 66.4 (59.0–73.7) | 11.4 (7.0–15.8) | 6.3 (1.2–11.3) | 10.4 (6.5–14.3) | 18.1 (11.9–24.3) | 70.8 (63.6–77.9) | 26.6 (19.9–33.4) | 74.0 (68.1–80.0) | 34.1 (26.7–41.5) | 6.8 (3.5–10.0) |
| NHPI-Multiple | 2.7 (0.0–5.6) | 24.5 (15.4–33.7) | 10.4 (3.3–17.5) | 8.1 (1.0–15.2) | 12.2 (4.9–19.6) | 11.7 (3.3–20.1) | 4.9 (0.7–9.2) | 20.0 (10.0–30.0) | 5.5 (0.6–10.4) | 7.7 (2.5–12.9) | 4.8 (0.6–9.1) | 13.2 (7.0–19.3) | 47.9 (36.8–58.9) | 19.5 (9.0–29.9) | 15.8 (6.6–25.1) | 18.8 (9.7–27.9) | 39.9 (30.5–49.3) | 7.0 (1.6–12.4) | 59.2 (47.5–70.9) | 12.6 (6.8–18.5) | 2.6 (0.0–5.5) | 16.5 (8.7–24.3) | 8.2 (2.7–13.8) | 58.9 (45.9–71.9) | 7.7 (3.1–12.3) | 71.6 (63.8–79.5) | 46.6 (33.6–59.5) | 5.7 (0.2–11.3) |
| NHPI-White | 0.5 (0.0–1.5) | 23.9 (13.7–34.1) | 7.6 (0.0–15.5) | 3.2 (0.0–8.7) | 17.1 (5.6–28.5) | 2.8 (0.0–8.2) | 3.3 (0.0–9.2) | 12.2 (3.0–21.5) | 0.0 (0.0–0.0) | 3.2 (0.0–6.5) | 8.1 (0.0–17.1) | 7.9 (0.5–15.3) | 58.7 (42.4–75.1) | 20.4 (6.7–34.2) | 16.2 (3.8–28.6) | 15.1 (3.7–26.4) | 38.5 (23.1–53.9) | 0.9 (0.0–2.1) | 54.2 (42.2–66.2) | 32.4 (22.9–41.9) | 0.5 (0.0–1.5) | 14.7 (5.5–23.9) | 10.3 (0.4–20.2) | 74.1 (65.1–83.2) | 8.5 (1.6–15.5) | 67.8 (56.3–79.4) | 45.1 (30.8–59.4) | 13.1 (2.7–23.5) |
| Native Hawaiian | 5.6 (0.0–12.3) | 28.5 (12.8–44.2) | 14.1 (4.5–23.6) | 2.3 (0.0–6.1) | 11.4 (2.4–20.4) | 2.6 (0.0–6.4) | 3.5 (0.0–8.0) | 10.3 (2.7–17.9) | 4.3 (0.0–11.7) | 2.7 (0.0–6.6) | 17.1 (6.6–27.6) | 15.2 (4.9–25.5) | 58.6 (43.8–73.3) | 19.6 (6.3–33.0) | 27.9 (15.8–40.0) | 15.2 (4.6–25.8) | 74.5 (61.9–87.0) | 2.3 (0.0–5.0) | 68.9 (56.9–80.9) | 4.6 (0.0–9.7) | 6.3 (0.0–12.9) | 7.4 (1.1–13.6) | 3.5 (0.0–8.2) | 58.4 (44.2–72.6) | 25.5 (16.4–34.5) | 63.7 (51.8–75.5) | 44.9 (31.3–58.4) | 0.7 (0.0–2.1) |
| Other Asian | 3.4 (1.5–5.3) | 6.1 (3.9–8.2) | 3.6 (1.9–5.3) | 0.5 (0.0–1.1) | 16.2 (11.9–20.5) | 2.1 (0.7–3.5) | 2.6 (0.8–4.4) | 10.4 (7.1–13.6) | 3.2 (1.4–4.9) | 4.4 (1.8–7.1) | 11.0 (7.7–14.2) | 9.5 (6.4–12.7) | 45.2 (40.5–50.0) | 9.4 (6.9–11.8) | 19.2 (15.4–23.0) | 15.4 (11.5–19.2) | 68.4 (63.7–73.2) | 11.0 (7.5–14.4) | 59.1 (54.1–64.1) | 7.0 (4.5–9.6) | 3.3 (1.6–4.9) | 8.3 (5.5–11.0) | 7.1 (4.6–9.7) | 50.6 (45.6–55.5) | 20.3 (16.0–24.6) | 72.2 (67.8–76.6) | 25.6 (21.7–29.5) | 3.8 (1.6–6.0) |
| Other Hispanic | 3.0 (1.9–4.1) | 11.3 (9.4–13.2) | 5.9 (4.5–7.4) | 1.0 (0.4–1.6) | 14.6 (12.6–16.6) | 2.5 (1.6–3.4) | 3.1 (2.1–4.1) | 14.0 (11.6–16.3) | 5.3 (3.8–6.7) | 8.6 (6.7–10.5) | 15.8 (13.5–18.1) | 15.1 (12.7–17.5) | 43.4 (40.2–46.6) | 14.3 (12.0–16.6) | 30.9 (27.8–33.9) | 29.0 (26.1–31.9) | 51.5 (48.4–54.5) | 16.7 (14.7–18.6) | 63.6 (60.6–66.7) | 12.2 (10.0–14.4) | 4.2 (2.8–5.6) | 11.4 (9.3–13.5) | 14.0 (11.8–16.3) | 75.5 (72.8–78.1) | 28.7 (26.2–31.2) | 74.2 (71.4–77.1) | 27.4 (24.6–30.2) | 9.1 (7.1–11.1) |
| Other Pacific Islander | 4.8 (0.1–9.4) | 11.8 (6.0–17.7) | 6.8 (1.7–12.0) | 0.0 (0.0–0.0) | 9.8 (4.1–15.5) | 2.1 (0.0–4.2) | 1.9 (0.1–3.8) | 15.5 (7.0–24.0) | 5.7 (0.1–11.3) | 3.8 (1.1–6.5) | 13.8 (6.2–21.4) | 24.2 (14.6–33.7) | 58.6 (49.2–68.0) | 19.3 (11.7–27.0) | 15.4 (8.5–22.3) | 18.6 (10.5–26.6) | 58.2 (48.1–68.4) | 5.9 (2.1–9.7) | 49.7 (40.2–59.3) | 7.4 (3.1–11.7) | 4.2 (0.1–8.3) | 12.0 (5.3–18.6) | 11.0 (3.8–18.2) | 72.2 (64.3–80.2) | 13.3 (7.9–18.6) | 67.8 (59.4–76.2) | 34.6 (25.4–43.8) | 7.3 (2.0–12.7) |
| Other Race | 3.4 (2.1–4.8) | 15.2 (12.5–17.9) | 7.8 (5.9–9.8) | 4.3 (3.0–5.5) | 15.3 (12.5–18.0) | 3.2 (1.5–4.8) | 5.7 (3.9–7.5) | 13.5 (10.8–16.1) | 3.8 (2.6–4.9) | 9.9 (7.6–12.2) | 16.6 (13.3–19.9) | 11.4 (8.8–14.1) | 55.5 (51.5–59.5) | 15.2 (12.7–17.8) | 18.0 (15.2–20.9) | 15.9 (13.1–18.6) | 55.1 (51.2–59.1) | 8.1 (6.2–10.0) | 67.0 (63.4–70.7) | 16.9 (13.9–19.9) | 3.1 (1.9–4.3) | 15.0 (12.3–17.7) | 12.4 (9.5–15.3) | 65.7 (62.0–69.4) | 22.8 (19.5–26.1) | 66.4 (62.7–70.2) | 38.6 (34.6–42.5) | 3.5 (2.2–4.8) |
| Other Race-White | 5.3 (0.0–11.1) | 18.3 (10.1–26.4) | 11.8 (4.9–18.8) | 7.4 (2.5–12.2) | 23.6 (13.2–33.9) | 5.7 (1.1–10.3) | 13.9 (5.5–22.3) | 17.1 (8.7–25.4) | 1.6 (0.0–3.4) | 13.7 (6.5–21.0) | 15.4 (8.6–22.2) | 10.6 (4.2–17.0) | 60.0 (48.7–71.4) | 20.4 (10.5–30.3) | 18.4 (9.2–27.6) | 21.1 (11.8–30.4) | 50.8 (39.5–62.2) | 11.4 (4.4–18.3) | 62.8 (52.4–73.3) | 23.9 (15.2–32.6) | 2.7 (0.0–6.2) | 20.3 (10.5–30.2) | 13.5 (6.0–21.1) | 65.7 (56.1–75.4) | 17.7 (9.9–25.6) | 69.7 (60.8–78.6) | 44.3 (32.9–55.7) | 10.8 (4.0–17.6) |
| Puerto Rican | 2.0 (0.0–4.3) | 12.4 (6.2–18.6) | 9.0 (3.5–14.6) | 0.0 (0.0–0.0) | 4.9 (1.9–8.0) | 3.0 (0.0–7.1) | 11.0 (3.3–18.7) | 6.2 (2.3–10.1) | 14.5 (6.4–22.6) | 16.2 (6.9–25.5) | 17.8 (9.7–26.0) | 9.0 (2.4–15.6) | 53.6 (42.3–64.8) | 11.0 (5.4–16.7) | 33.0 (20.8–45.2) | 23.6 (12.8–34.4) | 47.1 (35.7–58.6) | 4.0 (0.8–7.2) | 57.7 (46.3–69.1) | 10.6 (4.6–16.6) | 3.4 (0.0–7.1) | 8.5 (3.5–13.5) | 14.5 (5.9–23.1) | 72.1 (61.1–83.1) | 16.5 (8.9–24.1) | 63.0 (52.1–73.9) | 43.7 (32.8–54.5) | 4.0 (0.6–7.5) |
| Samoan | 3.0 (0.0–6.8) | 15.1 (5.1–25.2) | 7.6 (0.0–15.7) | 0.0 (0.0–0.0) | 17.1 (11.8–22.5) | 1.2 (0.0–3.4) | 12.5 (2.8–22.2) | 11.6 (3.0–20.1) | 2.0 (0.0–4.9) | 11.4 (1.7–21.2) | 8.8 (1.6–16.0) | 8.9 (0.0–18.1) | 42.8 (29.8–55.8) | 13.0 (2.8–23.1) | 15.6 (4.9–26.4) | 21.2 (10.0–32.4) | 45.2 (30.2–60.2) | 10.6 (2.0–19.2) | 75.9 (64.5–87.2) | 13.1 (3.6–22.6) | 4.0 (0.0–8.4) | 5.2 (0.2–10.3) | 6.1 (0.3–12.0) | 80.2 (68.3–92.0) | 22.6 (16.0–29.2) | 82.7 (72.3–93.0) | 46.6 (32.0–61.2) | 1.8 (0.0–4.4) |
| Unspecified Asian | 2.0 (0.4–3.6) | 11.7 (8.6–14.7) | 5.8 (3.4–8.1) | 1.1 (0.0–2.5) | 11.4 (8.5–14.3) | 2.6 (1.2–4.0) | 2.7 (1.1–4.4) | 7.7 (5.4–10.0) | 1.9 (0.5–3.4) | 3.5 (1.8–5.3) | 7.5 (5.1–10.0) | 12.5 (9.1–15.9) | 49.1 (44.2–53.9) | 9.0 (6.3–11.7) | 14.8 (11.4–18.1) | 11.9 (8.7–15.0) | 73.2 (69.1–77.3) | 2.3 (1.4–3.2) | 48.7 (43.9–53.5) | 8.9 (6.3–11.5) | 0.4 (0.0–0.8) | 10.7 (7.9–13.6) | 7.6 (5.0–10.3) | 42.8 (38.2–47.4) | 15.0 (12.1–17.9) | 70.5 (66.2–74.9) | 22.2 (18.1–26.2) | 3.5 (1.4–5.5) |
| Unspecified NHPI | 6.3 (0.0–12.6) | 14.4 (7.1–21.7) | 8.4 (3.0–13.9) | 4.0 (0.0–8.4) | 9.3 (3.2–15.3) | 0.3 (0.0–1.0) | 6.8 (1.7–12.0) | 8.2 (0.3–16.1) | 1.3 (0.0–3.1) | 12.3 (2.0–22.5) | 7.1 (0.7–13.4) | 21.6 (11.4–31.9) | 54.7 (41.9–67.5) | 17.0 (7.7–26.3) | 18.5 (9.2–27.7) | 5.1 (0.7–9.5) | 61.1 (50.1–72.2) | 9.7 (0.7–18.6) | 58.2 (46.7–69.8) | 11.3 (2.1–20.5) | 1.1 (0.0–3.0) | 8.2 (0.7–15.8) | 7.5 (1.6–13.4) | 69.9 (57.9–81.8) | 19.6 (10.2–28.9) | 77.6 (69.2–86.0) | 39.3 (26.9–51.7) | 2.6 (0.0–5.3) |
| Vietnamese | 1.8 (0.1–3.4) | 10.8 (6.4–15.2) | 7.5 (3.4–11.6) | 0.0 (0.0–0.0) | 10.6 (7.6–13.6) | 0.9 (0.0–1.9) | 2.1 (0.2–4.1) | 6.5 (3.5–9.6) | 4.0 (1.0–7.1) | 1.5 (0.3–2.6) | 5.2 (2.1–8.4) | 6.1 (2.8–9.3) | 40.8 (34.8–46.8) | 9.6 (6.8–12.5) | 15.3 (10.8–19.7) | 11.8 (7.1–16.4) | 60.4 (53.1–67.8) | 4.6 (2.0–7.1) | 47.7 (40.3–55.0) | 8.0 (4.8–11.2) | 3.2 (0.9–5.5) | 6.3 (2.8–9.9) | 3.7 (1.6–5.9) | 32.0 (25.8–38.2) | 12.4 (8.8–16.0) | 74.9 (68.6–81.2) | 21.1 (16.6–25.5) | 5.6 (0.2–10.9) |
| White | 3.5 (3.3–3.7) | 16.5 (15.9–17.0) | 9.8 (9.4–10.2) | 9.6 (9.2–10.0) | 16.3 (15.8–16.8) | 2.9 (2.7–3.1) | 5.5 (5.3–5.8) | 10.0 (9.6–10.4) | 3.1 (2.8–3.3) | 6.1 (5.8–6.4) | 10.7 (10.3–11.1) | 7.5 (7.2–7.8) | 64.3 (63.6–64.9) | 19.0 (18.5–19.6) | 16.1 (15.6–16.6) | 11.9 (11.5–12.3) | 53.1 (52.4–53.7) | 4.8 (4.5–5.1) | 57.4 (56.7–58.0) | 20.0 (19.5–20.5) | 3.4 (3.2–3.6) | 12.8 (12.3–13.2) | 10.4 (10.0–10.8) | 61.5 (60.8–62.1) | 17.8 (17.3–18.3) | 67.0 (66.4–67.5) | 41.0 (40.4–41.7) | 3.3 (3.1–3.5) |

Abbreviations: BRFSS = Behavioral Risk Factor Surveillance System; HIV = human immunodeficiency virus; COPD = chronic obstructive pulmonary disorder

^a^ Reported figures are unweighted N and weighted proportions from multiply imputed data (20 imputations, 20 iterations)

**Appendix C.** Distribution of health indicators in study population, overall and stratified by sex and by age group, California BRFSS 2014-2023

|  | **Overall**  100,177 (100%) | **Female**  50,992 (51%) | **Male**  49,185 (49%) | **18 to 29 years**  17,580 (22%) | **30 to 44 years**  23,496 (27%) | **45 to 59 years**  24,194 (24%) | **60 years and over**  34,907 (27%) |
| --- | --- | --- | --- | --- | --- | --- | --- |
| Lifestyle factors |  |  |  |  |  |  |  |
| Ever smoked | 35,018 (32.5%) | 15,156 (25.8%) | 19,862 (39.4%) | 3,361 (18.0%) | 7,266 (30.8%) | 8,930 (35.4%) | 15,461 (43.1%) |
| Any alcohol use | 56,253 (53.7%) | 25,702 (47.5%) | 30,551 (60.2%) | 10,589 (55.5%) | 14,109 (57.3%) | 13,671 (54.7%) | 17,884 (47.9%) |
| Binge drinking | 15,989 (16.4%) | 5,804 (11.6%) | 10,185 (21.3%) | 4,901 (25.5%) | 5,108 (20.5%) | 3,662 (14.6%) | 2,318 (6.6%) |
| No exercise in last 30 days | 20,526 (21.2%) | 11,175 (22.4%) | 9,351 (20.0%) | 2,807 (17.9%) | 4,432 (20.1%) | 5,245 (22.1%) | 8,042 (24.3%) |
| Preventive healthcare |  |  |  |  |  |  |  |
| Never had HIV test | 58,242 (58.8%) | 29,573 (58.3%) | 28,669 (59.2%) | 10,634 (62.6%) | 9,942 (44.9%) | 12,075 (52.5%) | 25,591 (75.0%) |
| No flu vaccine in last 12 months | 57,765 (60.1%) | 27,980 (57.1%) | 29,785 (63.2%) | 12,475 (71.5%) | 15,780 (67.8%) | 15,052 (61.1%) | 14,458 (42.5%) |
| Never had pneumonia vaccine | 63,858 (67.8%) | 31,287 (66.1%) | 32,571 (69.6%) | 12,644 (72.1%) | 19,202 (82.5%) | 18,728 (78.3%) | 13,284 (40.6%) |
| No routine checkup in last 2 years | 14,934 (15.5%) | 5,983 (12.1%) | 8,951 (19.0%) | 3,864 (20.9%) | 4,985 (20.8%) | 3,667 (14.4%) | 2,418 (6.8%) |
| No primary care provider | 21,900 (23.5%) | 8,642 (18.7%) | 13,258 (28.5%) | 7,442 (40.8%) | 7,464 (31.0%) | 4,429 (17.8%) | 2,565 (7.4%) |
| No health insurance | 9,357 (10.6%) | 3,996 (9.0%) | 5,361 (12.2%) | 2,368 (14.5%) | 3,449 (15.0%) | 2,537 (10.5%) | 1,003 (3.2%) |
| Chronic conditions |  |  |  |  |  |  |  |
| Angina or coronary heart disease | 3,879 (3.2%) | 1,698 (2.6%) | 2,181 (3.8%) | 63 (0.4%) | 167 (0.8%) | 654 (2.6%) | 2,995 (8.2%) |
| Heart attack | 3,887 (3.2%) | 1,647 (2.5%) | 2,240 (3.9%) | 55 (0.4%) | 228 (1.0%) | 794 (3.2%) | 2,810 (7.6%) |
| Diabetes diagnosis | 11,077 (10.6%) | 5,608 (10.4%) | 5,469 (10.8%) | 180 (1.1%) | 907 (4.3%) | 3,038 (13.0%) | 6,952 (22.2%) |
| Chronic kidney disease | 3,385 (2.9%) | 1,876 (3.0%) | 1,509 (2.7%) | 131 (0.7%) | 331 (1.4%) | 722 (2.9%) | 2,201 (6.0%) |
| Lifetime asthma | 14,852 (14.2%) | 8,561 (15.9%) | 6,291 (12.4%) | 2,871 (16.0%) | 3,341 (13.5%) | 3,471 (13.4%) | 5,169 (14.1%) |
| Current asthma | 8,761 (8.2%) | 5,546 (10.2%) | 3,215 (6.2%) | 1,379 (7.9%) | 1,807 (7.5%) | 2,243 (8.5%) | 3,332 (9.1%) |
| COPD | 5,556 (4.6%) | 3,258 (5.0%) | 2,298 (4.2%) | 279 (1.6%) | 514 (2.2%) | 1,247 (4.3%) | 3,516 (9.5%) |
| Skin cancer | 7,613 (5.2%) | 4,079 (5.4%) | 3,534 (5.0%) | 59 (0.4%) | 241 (1.0%) | 1,184 (4.0%) | 6,129 (14.3%) |
| Overweight or obese | 62,468 (62.3%) | 29,127 (56.8%) | 33,341 (68.0%) | 8,190 (46.9%) | 15,054 (65.6%) | 16,925 (69.9%) | 22,299 (64.5%) |
| Major depression | 16,888 (15.2%) | 10,692 (19.0%) | 6,196 (11.3%) | 2,770 (15.6%) | 3,771 (14.7%) | 4,382 (15.6%) | 5,965 (15.2%) |
| General health status |  |  |  |  |  |  |  |
| Fair or poor health status | 18,236 (17.8%) | 9,893 (18.8%) | 8,343 (16.8%) | 1,601 (9.7%) | 3,164 (14.3%) | 5,262 (21.3%) | 8,209 (24.6%) |
| Frequent poor physical health | 11,582 (10.4%) | 6,552 (11.3%) | 5,030 (9.5%) | 824 (4.9%) | 1,784 (7.7%) | 3,346 (12.6%) | 5,628 (15.4%) |
| Frequent poor mental health | 11,583 (11.5%) | 6,604 (12.9%) | 4,979 (10.1%) | 2,412 (14.0%) | 2,820 (11.9%) | 3,050 (11.5%) | 3,301 (9.2%) |
| Disabilities |  |  |  |  |  |  |  |
| Vision impairment | 4,878 (4.8%) | 2,791 (5.3%) | 2,087 (4.2%) | 469 (2.9%) | 621 (3.0%) | 1,455 (5.9%) | 2,333 (7.1%) |
| Difficulty walking | 13,611 (11.7%) | 8,190 (13.6%) | 5,421 (9.8%) | 342 (2.2%) | 1,165 (5.3%) | 3,414 (12.9%) | 8,690 (24.6%) |
| Cognitive difficulty | 10,824 (10.8%) | 5,997 (11.6%) | 4,827 (10.1%) | 2,114 (12.9%) | 2,246 (9.7%) | 2,847 (10.5%) | 3,617 (10.6%) |
| Difficulty running errands | 6,786 (6.3%) | 4,229 (7.5%) | 2,557 (5.1%) | 681 (4.3%) | 997 (4.5%) | 1,769 (6.6%) | 3,339 (9.5%) |
| Difficulty dressing | 4,084 (3.6%) | 2,271 (3.7%) | 1,813 (3.5%) | 130 (1.0%) | 463 (2.1%) | 1,246 (4.6%) | 2,245 (6.3%) |
